# T-cell mediated immunity after AZD1222 vaccination: A polyfunctional spike-specific Th1 response with a diverse TCR repertoire

**DOI:** 10.1101/2021.06.17.21259027

**Authors:** Phillip A. Swanson, Marcelino Padilla, Wesley Hoyland, Kelly McGlinchey, Paul A. Fields, Sagida Bibi, Saul N. Faust, Adrian B. McDermott, Teresa Lambe, Andrew J. Pollard, Nicholas M. Durham, Elizabeth J. Kelly

## Abstract

AZD1222 (ChAdOx1 nCoV-19), a replication-deficient simian adenovirus-vectored vaccine, has demonstrated safety, efficacy, and immunogenicity against coronavirus disease 2019 (COVID-19) in clinical trials and real-world studies. We characterized CD4+ and CD8+ T-cell responses induced by AZD1222 vaccination in peripheral blood mononuclear cells (PBMCs) from 280 unique vaccine recipients aged 18–85 years who enrolled in the phase 2/3 COV002 trial. Total spike-specific CD4+ T cell helper type 1 (Th1) and CD8+ T-cell responses were significantly increased in AZD1222-vaccinated adults of all ages following two doses of AZD1222. CD4+ Th2 responses following AZD1222 vaccination were not detected. Furthermore, AZD1222-specific Th1 and CD8+ T cells both displayed a high degree of polyfunctionality in all adult age groups. T-cell receptor (TCR) β sequences from vaccinated participants mapped against TCR sequences known to react to SARS-CoV-2 revealed substantial breadth and depth across the SARS-CoV-2 spike protein for the AZD1222-induced CD4+ and CD8+ T-cell responses. Overall, AZD1222 vaccination induced a robust, polyfunctional Th1-dominated T-cell response, with broad CD4+ and CD8+ T-cell coverage across the SARS-CoV-2 spike protein.

**One Sentence Summary:** Polyfunctional CD4+ and CD8+ T-cell responses are elicited against the SARS-CoV-2 spike protein following vaccination with AZD1222

## Introduction

The coronavirus disease 2019 (COVID-19) pandemic caused by severe acute respiratory syndrome coronavirus 2 (SARS-CoV-2) is continuing to cause substantial widespread morbidity and mortality worldwide (*1*). Cellular immunity may be critical in an individual’s response to SARS-CoV-2 infection, particularly in the face of neutralizing-antibody (nAB) escape by emerging variants, and for reducing the severity of COVID-19 disease. Cases of asymptomatic COVID-19 have been associated with T-cell responses without seroconversion, suggesting a role for T-cell responses following SARS-CoV-2 exposure in the absence of nAbs (*2*). Furthermore, studies of patients with COVID-19 show that the presence of SARS-CoV-2-specific CD4+ and CD8+ T cells are associated with lower disease severity (*3–5*). SARS-CoV-2 vaccines have also been shown to elicit T-cell responses (*6–10*), but more in-depth analyses of the functionality and breadth of vaccine-specific T cells could provide valuable information on potential determinants of protection.

AZD1222 (ChAdOx1 nCoV-19) is a replication-deficient simian adenovirus-vectored vaccine indicated for the prevention of COVID-19. The safety, efficacy, and immunogenicity of AZD1222 has been extensively demonstrated in a global clinical development program, supporting regulatory submissions for conditional or emergency use of AZD1222 (*11–14*). In a phase 1/2 trial conducted in the UK, COV001, AZD1222 induced marked increases in SARS-CoV-2 spike-specific effector T-cell responses in adults aged 18–55 years, which peaked at Day 14 post vaccination. These responses were reported as early as Day 8 and were maintained through Day 56, which was the latest timepoint analyzed (*12*). Similar responses were reported in adults aged 18 to ≥70 years in a phase 2/3 study conducted in the UK, COV002, in which SARS-CoV-2 spike-specific effector T-cell responses peaked at Day 14 post vaccination (*13*).

Peak responses in adults 18–55 years old were characterized by IFNγ-, IL-2-, and TNF-producing T cell helper type 1 (Th1) CD4+ T cells and CD8+ T cells (*11*). However, the cytokine profile of AZD1222-induced T-cell responses beyond Day 14 in younger adults or following a second dose of AZD1222 remains unknown. Furthermore, a full characterization of AZD1222-induced T-cell cytokine responses in adults older than 55 is also undetermined (*11*).

Here, we aimed to characterize functional CD4+ and CD8+ T-cell responses following the first and second doses of AZD1222 in healthy adults aged 18–85 years enrolled in phase 1–3 trials conducted in the UK. Furthermore, we describe the breadth and depth of unique SARS-CoV-2 spike-specific T-cell responses induced by AZD1222 vaccination. These data provide a comprehensive analysis of the AZD1222 T-cell response in an adult population.

## Results

### Study participants

Evaluable samples were obtained from 280 participants enrolled in the COV001 and COV002 studies. Participant samples analyzed in this study were selected from three different age cohorts who received two doses of either 5×10^10^ virus particles of AZD1222 or control meningococcal conjugate vaccine (MenACWY) (Table 1). Of the 280 participants, 67 participants were analyzed for spike-specific cytokine secretion by intracellular cytokine staining (ICS) and 233 were analyzed by T-cell receptor (TCR) sequencing. Twenty participants had samples in both sets of analyses.

**Table 1.**
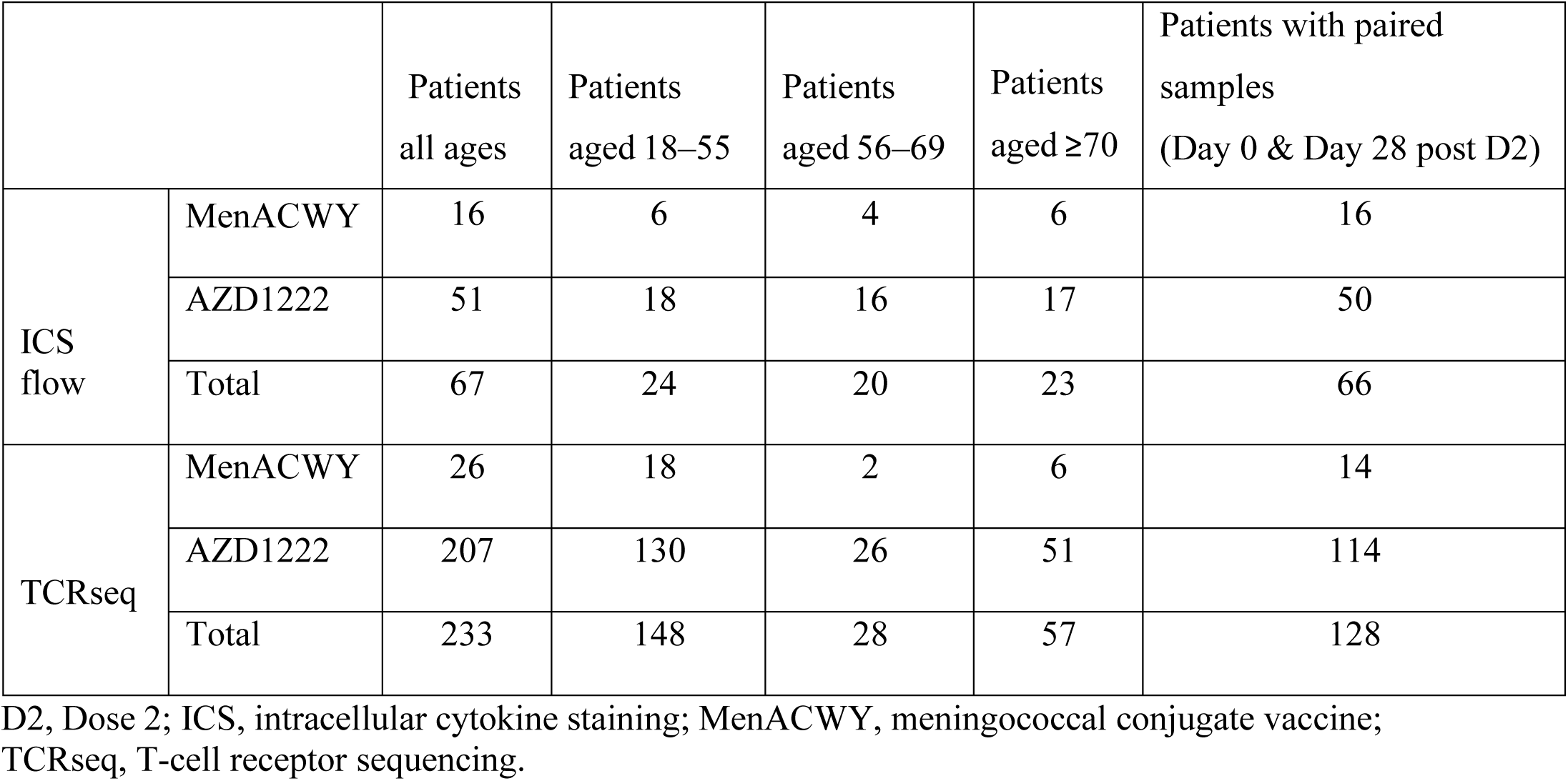
Details of participants included in this analysis.

|  |  | Patients<br>all ages | Patients<br>aged 18–55 | Patients<br>aged 56–69 | Patients<br>aged ≥70 | Patients with paired<br>samples<br>(Day 0 & Day 28 post D2) |
| --- | --- | --- | --- | --- | --- | --- |
| ICS<br>flow | MenACWY | 16 | 6 | 4 | 6 | 16 |
|  | AZD1222 | 51 | 18 | 16 | 17 | 50 |
|  | Total | 67 | 24 | 20 | 23 | 66 |
| TCRseq | MenACWY | 26 | 18 | 2 | 6 | 14 |
|  | AZD1222 | 207 | 130 | 26 | 51 | 114 |
|  | Total | 233 | 148 | 28 | 57 | 128 |

D2, Dose 2; ICS, intracellular cytokine staining; MenACWY, meningococcal conjugate vaccine; TCRseq, T-cell receptor sequencing.

### AZD1222 induced strong Th1-biased T-cell responses to the SARS-CoV-2 spike protein

SARS-CoV-2 spike-specific T-cell responses from all participants vaccinated with AZD1222 or MenACWY were assessed by measuring intracellular cytokine production in peripheral blood mononuclear cells (PBMCs) following in vitro stimulation with two separate overlapping peptide pools covering the entire SARS-CoV-2 spike protein sequence.

Total spike-specific Th1 responses significantly increased in AZD1222-vaccinated participants at Day 28 post vaccination and at Day 56 post vaccination (4 weeks following the second dose), compared with Day 0 (Fig. 1A). In MenACWY-vaccinated participants, no increases in spike-specific Th1 responses were observed over time. Th1 responses in AZD1222-vaccinated participants were significantly greater than those who received MenACWY, at both Day 28 and Day 56 post vaccination. The Th1 response followed a hierarchical pattern of cytokine production, with the majority primarily producing TNF, followed by IL-2 and IFNγ at both Day 28 and Day 56 post vaccination (Fig. 1B and fig. S1A). Spike-specific Th2 responses were near absent following AZD1222 vaccination (Fig. 1C–D and fig. S1B).

**Fig. 1.**
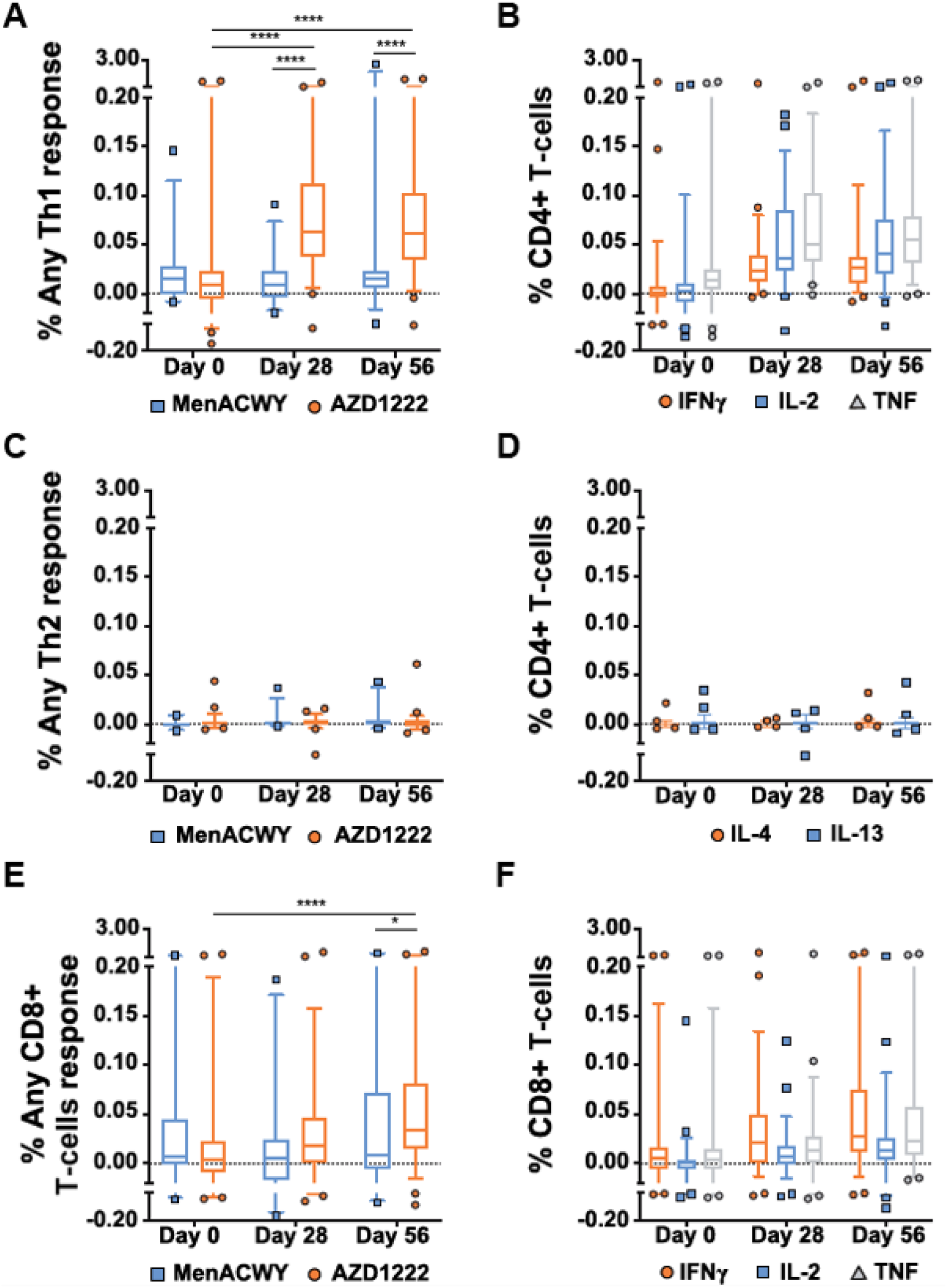
T-cell responses following vaccination with AZD1222 or MenACWY. PBMCs from human participants vaccinated with AZD1222 or MenACWY at the indicated timepoint post vaccination were stimulated with SARS-CoV-2 spike peptide pools, and the intracellular cytokine response was measured. (A) Frequencies of total spike-specific CD4+ T cells producing any combination of Th1 (IFNγ, IL-2, or TNF) or (C) Th2 (IL-4 or IL-13) cytokines. (B) Frequencies of individual Th1 or (D) Th2 cytokines at the indicated timepoints in participants vaccinated with AZD1222. (E) Frequencies of total spike-specific CD8+ T cells producing any combination of IFNγ, IL-2, or TNF following vaccination with AZD1222 or MenACWY. (F) Frequencies of individual CD8+ T-cell cytokines at the indicated timepoints in participants vaccinated with AZD1222. For all data, responses to each peptide pool were combined to determine the total spike-specific response. In the box and whisker plots, the horizontal line represents median, boxes represent interquartile range, whiskers extend to the 5th and 95th percentiles, and symbols represent outlier samples. Significant differences between AZD1222 and MenACWY at each timepoint were determined by two-tailed Mann-Whitney tests. Significant differences between timepoints within each vaccine group were determined by Kruskal-Wallis test with Dunn’s test to correct for multiple comparisons. *p<0.05, ****p<0.0001. MenACWY, meningococcal conjugate vaccine; PBMCs, peripheral blood mononuclear cells; SARS-CoV-2, severe acute respiratory syndrome coronavirus 2; Th1, T cell helper type 1; Th2, T cell helper type 2.

In comparison with the Th1 response, total spike-specific CD8+ T cells were detected at lower frequencies at Day 28 and Day 56 post vaccination. However, at Day 56 post AZD1222 vaccination, spike-specific CD8+ T cells were significantly elevated compared with Day 0 and compared with Day 56 responses in participants vaccinated with MenACWY (Fig. 1E). Spike-specific CD8+ T cells were distinct from CD4+ T cells, as the majority primarily produced IFNγ, followed by TNF and IL-2 (Fig. 1F and fig. S1C). These data demonstrate that AZD1222 elicited a robust Th1-dominated T-cell response with a significantly expanded CD8+ T-cell response post vaccination compared with Day 0, albeit at a smaller magnitude than the CD4+ response.

### AZD1222 vaccination generated a polyfunctional CD4+ T-cell cytokine response across adult age groups

To determine if CD4+ T-cell responses to AZD1222 vaccination varied by age, subgroup analyses according to participants’ age, 18–55, 56–69, and ≥70 years, were performed. Significant increases in spike-specific CD4+ T-cell responses over Day 0 were observed for participants within each age group, with a similar pattern of Th1 cytokine hierarchy (TNF>IL-2>IFNγ) (Fig. 2A and fig. S2A). However, the kinetics of the AZD1222-specific Th1 cell response differed by age. In participants aged 18–55 years, spike-specific Th1 cells were elevated at Day 28 and remained at a consistent frequency through Day 56 post vaccination (Fig. 2A). In participants aged 55–69 years, responses peaked at Day 28 and declined by Day 56, despite receiving a second vaccine dose. Although Th1 cell responses observed at Day 28 post vaccination were at a lower frequency in participants aged ≥70 years compared with other age groups, spike-specific responses demonstrated an increased frequency at Day 56, potentially aided by the second vaccine dose at Day 28. Importantly, no age-specific differences in AZD1222-specific Th1 cell responses were observed at Day 56.

**Fig. 2.**
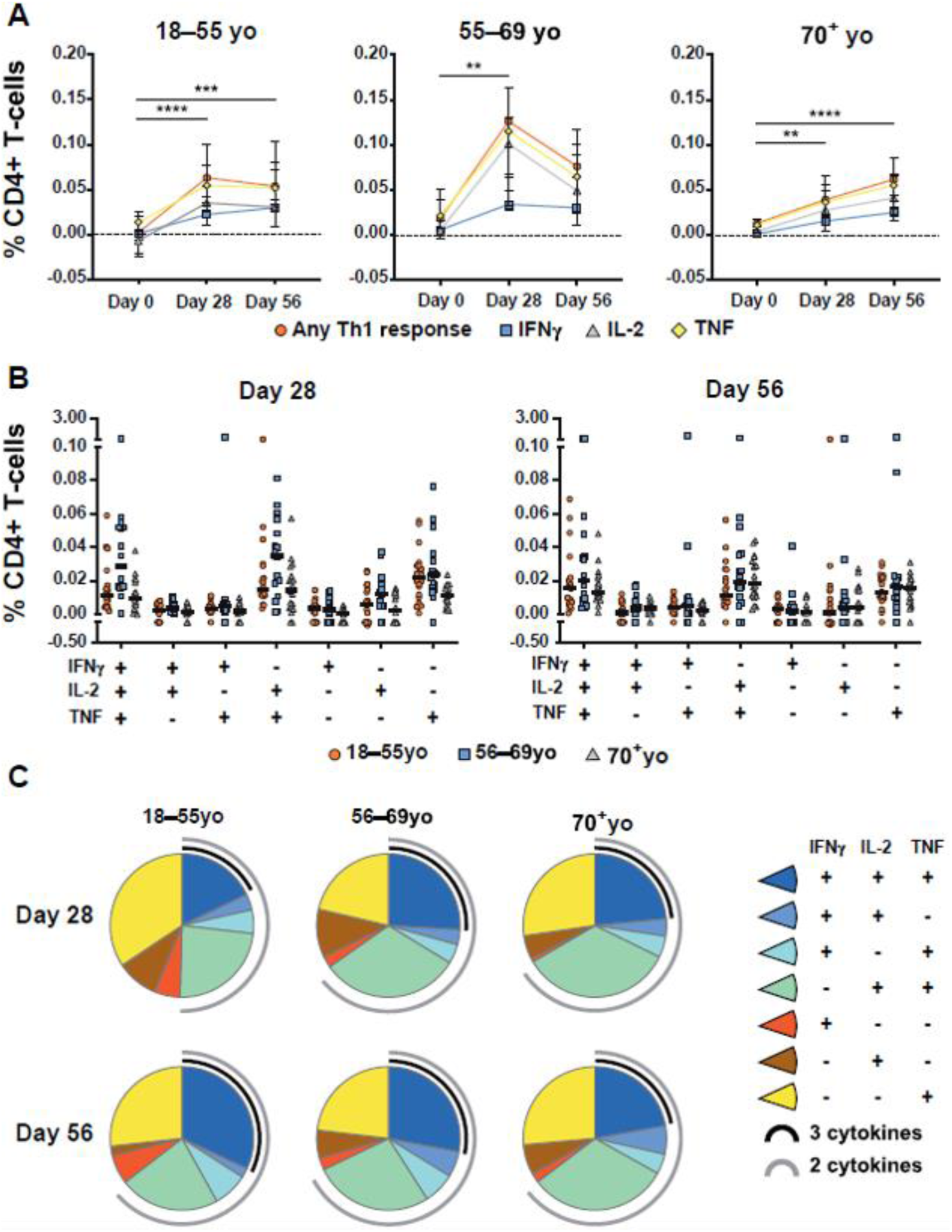
Age-specific CD4+ T-cell responses to AZD1222 vaccination. (A) Median frequencies with interquartile ranges of CD4+ T cells from participants within each age cohort producing IFNγ, IL-2, TNF, or any combination of these cytokines at the indicated timepoints following stimulation with SARS-CoV-2 spike peptide pools. Significant differences between timepoints within each vaccine group were determined by Kruskal-Wallis tests with Dunn’s test to correct for multiple comparisons. **p<0.01, ***p<0.001, ****p<0.0001. (B) Frequencies of antigen-stimulated CD4+ T cells producing each combination of IFNγ, IL-2, TNF cytokines at Day 28 (left) or Day 56 (right) post vaccination. Individual participant responses are shown with median represented by the horizontal line. (C) Pie graphs indicating the total proportion of spike-specific Th1 cytokine production averaged for all participants within the indicated age groups at Day 28 and Day 56 post vaccination. Proportion of multicytokine responses are represented by the black (three cytokines) and gray (two cytokines) arcs.

The frequencies (Fig. 2B) and proportions (Fig. 2C) of AZD1222-specific Th1 cells producing each combination of Th1 cytokines were compared among age groups at Day 28 and Day 56 post vaccination. Spike-specific Th1 cells displayed a high degree of polyfunctionality at Day 28 post vaccination in all three age groups, with only minimal differences in the proportion of cytokine-producing cells observed through Day 56 among participants aged 56–69 and ≥70 years. However, in participants aged 18–55 years, the proportion of polyfunctional spike-specific Th1 cells increased through Day 56. At Day 56, two-thirds of all spike-specific Th1 cells across all three cohorts were polyfunctional, with >25% of spike-specific Th1 cells in participants aged 18–55 and 56–69 years and nearly 25% in participants aged ≥70 years producing all three cytokines. These data show that AZD1222 induces a polyfunctional Th1 cell response that is equivalent in frequency and functionality across all adult age groups at Day 56 post vaccination.

### Polyfunctional CD8+ T-cell responses are generated following AZD1222 vaccination

AZD1222-specific CD8+ T cell responses were generated in all age groups, but at lower frequencies compared with the Th1 responses. Spike-specific CD8+ T-cell responses in participants aged 18–55 and 56–69 years peaked at Day 28 post vaccination, with no measurable effect at Day 56 post initial vaccination following a second dose at Day 28 (Fig. 3A and fig. S2B). However, participants aged ≥70 years seemed to benefit from a second dose, as significant expansion of spike-specific CD8+ T cells was not observed until Day 56 post vaccination.

**Fig. 3.**
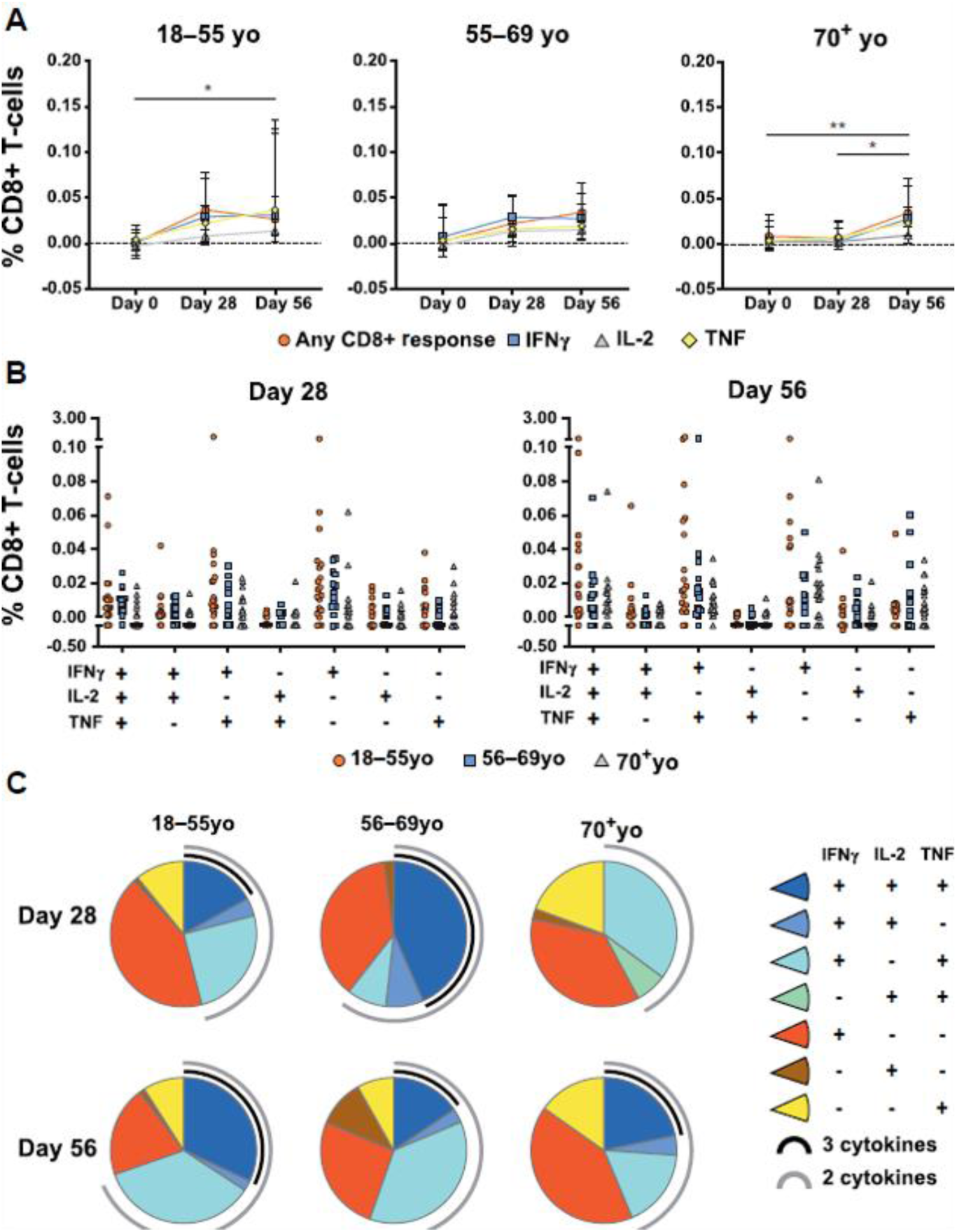
Age-specific CD8+ T-cell responses to AZD1222 vaccination. Median frequencies with interquartile ranges of CD8+ T cells producing IFNγ, IL-2, TNF, or any combination of these cytokines at the indicated timepoints following stimulation with SARS-CoV-2 spike peptide pools. Each symbol represents a participant within the indicated age cohort. Significant differences between timepoints within each vaccine group were determined by Kruskal-Wallis tests with Dunn’s test to correct for multiple comparisons. *p<0.05, **p<0.01. (A) Frequencies of antigen-stimulated CD8+ T cells producing each combination of IFNγ, IL-2, TNF cytokines at Day 28 (left) or Day 56 (right) post vaccination at the individual level (each symbol=individual participant). (C) Pie graphs indicating the total proportion of spike-specific CD8 T-cell cytokine production within the indicated age groups at Day 28 and Day 56 post vaccination. Proportion of multi-cytokine responses are represented by the black (three cytokines) and gray (two cytokines) arcs.

Importantly, by Day 56, no statistical differences in spike-specific CD8+ T-cell responses were observed among age groups.

The frequency and proportion of each combination of spike-specific CD8+ T-cell cytokines was also examined for each age group. At Day 28 post vaccination, most responding CD8+ T cells in participants aged 18–55 and ≥70 years only produced a single cytokine (Fig. 3B–C). However, participants aged 56–69 years generated a spike-specific CD8+ T-cell multicytokine response at Day 28 post vaccination. Although the total frequency of spike-specific CD8+ T cells in participants aged 18–55 years did not increase from Day 28 through Day 56, the proportion of multicytokine-producing cells increased from a minority to nearly 75% of all responding CD8+ T cells, with nearly one-third of responding CD8+ T cells producing TNF, IL-2, and IFNγ. Frequencies of polyfunctional, spike-specific CD8+ T cells in participants aged ≥70 years also increased from Day 28 through Day 56, with a large proportion producing TNF, IL-2, and IFNγ at Day 56. Together, these data demonstrate that AZD1222-specific CD8+ T cells are largely polyfunctional, and that the size of the response is equivalent across all age groups at Day 56 post vaccination.

### AZD1222-specific T-cell responses were genetically diverse and covered a broad range of SARS-CoV-2 spike epitopes

To better understand the diversity and specificity of the T-cell response to SARS-CoV-2 spike protein following AZD1222 vaccination, TCRβ chain sequencing was performed on 233 PBMC samples collected at Day 0 and Day 28 post second dose. Individual TCRs in each repertoire were compared against a library of TCRs with known specificity for SARS-CoV-2 that had been generated using the Multiplex Identification of T-cell Receptor Antigen Specificity (MIRA) platform (*15*). The total number of unique and total antigen-specific TCRs pre and post vaccination were characterized for each repertoire. PBMCs from study participants treated with two doses of AZD1222 at different dose intervals were used to increase the power of the TCR repertoire analysis. In addition to the participants used in the ICS analysis who received a second dose at approximately 4 weeks (18–60 days) post initial vaccination, a second cohort of participants who received a second dose at approximately 12 weeks (61–130 days) post vaccination were also analyzed. Participants vaccinated with AZD1222 at approximately 4-week and approximately 12-week second dose schedules both had a significant increase in the fraction of total TCRs that were spike-specific (depth) (fig. S3A) and fraction of unique TCRs that were spike-specific (breadth) (fig. S3B) post second dose. There was no statistical difference in either the depth or the breadth of the responses between the two dosing regimens. Because of this equivalency, data from these two dosing regimens were combined into single “Day 0” and “Day 28 post second dose” groups.

Looking first at the breadth of spike-specific TCRs from AZD1222-vaccinated participants, a significant increase was observed from Day 0 to Day 28 post second dose (Fig. 4A). Furthermore, spike-specific TCR breadth was also significantly elevated in AZD1222-vaccinated participants compared with MenACWY-vaccinated participants at Day 28 post second dose. No increase in TCR breadth of non-spike-specific SARS-CoV-2 TCRs was observed in either AZD1222- or MenACWY-vaccinated participants (fig. S4A). When participants were separated by age cohorts, all three age groups had a significant increase in spike-specific TCR breadth from Day 0 to Day 28 post second dose (Fig. 4B). In addition to TCR breadth, total TCR templates were also quantified for each group. Further confirming the ICS data (Fig. 1), the number of spike-specific TCRs, or depth, was significantly increased in AZD1222-vaccinated participants at Day 28 post second dose compared with Day 0, and also compared with MenACWY-vaccinated participants (Fig. 4C). There were no changes in depth within non– spike-specific TCRs in AZD1222- or MenACWY-vaccinated participants (fig. S4B). Participants aged 18–55 years and ≥70 years displayed significant increases in spike-specific TCR depth post vaccination (Fig. 4D).

**Fig. 4.**
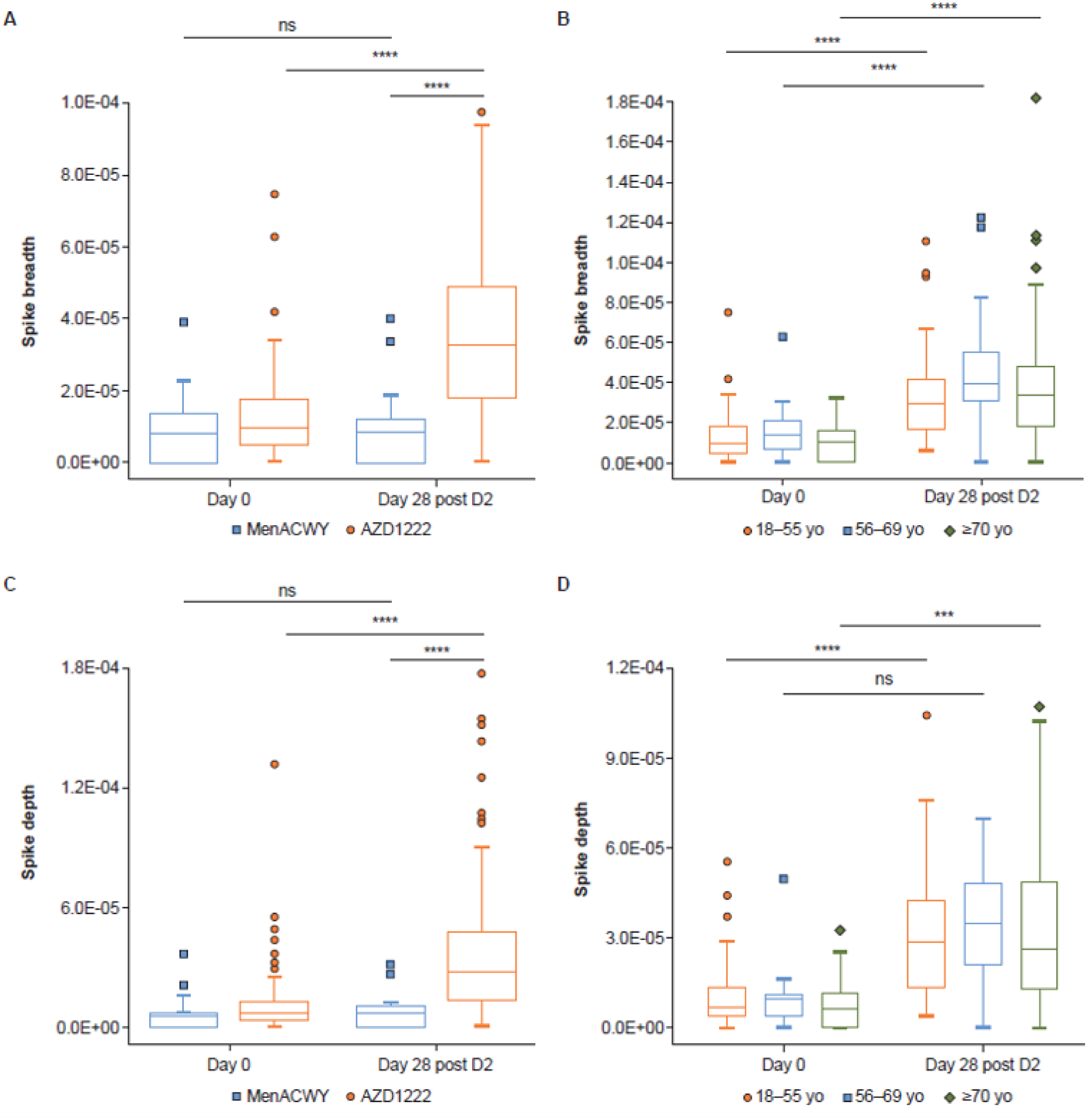
Spike-specific TCR breadth and depth following vaccination with AZD1222 or MenACWY. (A) Spike-specific TCR breadth following vaccination with MenACWY (blue) or AZD1222 (orange). (B) Spike-specific TCR breadth by age. (C) Spike-specific TCR depth following vaccination with MenACWY (blue) or AZD1222 (orange). (D) Spike-specific TCR depth by age. In the box and whisker plots, the horizontal line represents median, boxes represent interquartile range, whiskers extend to 5th and 95th percentiles, and symbols represent outlier samples. Significant differences determined by Sidak’s multiple comparisons tests. ns, not significant, ***p<0.001, ****p<0.0001. Breadth, SARS-CoV-2 associated unique TCRs/total unique TCRs; D2, Dose 2; Depth, SARS-CoV-2 associated T cells/total T cells; MenACWY, meningococcal conjugate vaccine; SARS-CoV-2, severe acute respiratory syndrome coronavirus 2.

To better understand the breadth and depth of the T-cell response in AZD1222-vaccinated participants, each unique spike-specific TCR sequence was mapped to the region of the spike protein to which it reacted in the MIRA data (*15*). Spike-specific CD4+ T-cell responses spanned the entire spike protein (Fig. 5A,B). Dominant responses were observed in the N-terminal region 160–218 and the C-terminal side 743–854. Significantly fewer unique spike-specific CD8+ T-cell TCRs were sequenced, likely due to the lower response post vaccination at these timepoints (Fig. 1) and the greater human leukocyte antigen (HLA) restriction of class I peptides. Half of all unique CD8+ TCRs mapped to a single region of the spike protein, with the other TCRs spread evenly across the other regions (Fig. 5C,D). Overlaying known mutations in variants of concern, including B.1.1.7 and B.1.351, indicates that most of the dominant responses are in regions not impacted by either variant. Therefore, these data demonstrate that AZD1222 vaccination induces a significant expansion of SARS-CoV-2 spike-specific T cells with TCR sequences that map to a number of epitopes across the entire spike protein.

**Fig. 5.**
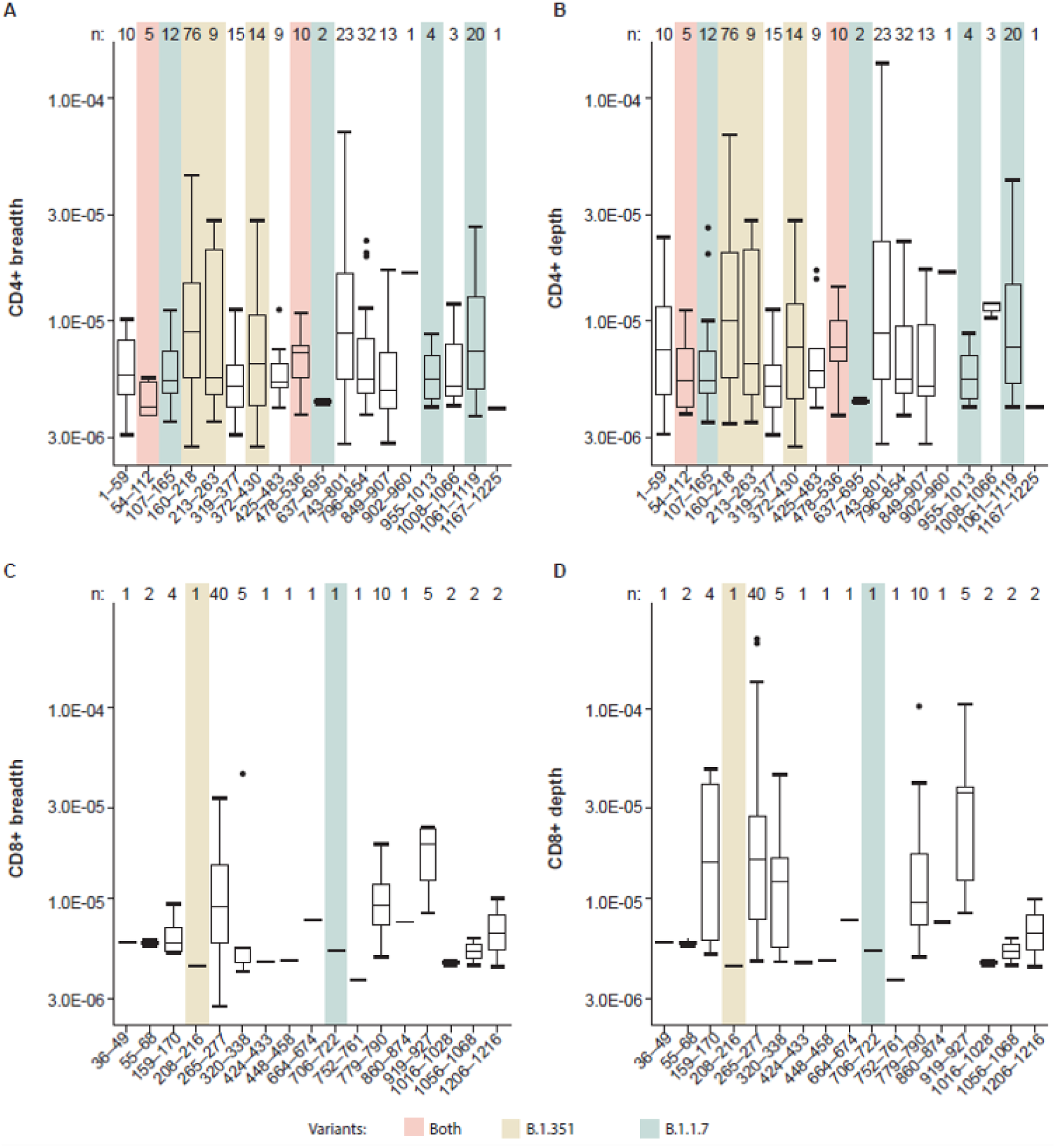
Spike-specific CD4+ and CD8+ T-cell breadth and depth at Day 28 post second dose. TCR sequences from participants vaccinated with AZD1222 at Day 28 post second dose were mapped against TCR sequences known to react to SARS-CoV-2. CD4+ breadth (A) and depth (B), and CD8+ breadth (C) and depth (D) were analyzed for each participant. The number of participants displaying responses for each epitope region is indicated (n). Sequences of known variants were aligned to known MIRA antigen locations, with any mutations observed in B.1.351 (yellow), B.1.1.7 (green), or both (red) highlighted. In the box and whisker plots, the horizontal line represents median, boxes represent interquartile range, whiskers extend to 5th and 95th percentiles, and symbols represent outlier samples. Breadth, SARS-CoV-2-associated unique TCRs/total unique TCRs; Depth, SARS-CoV-2-associated T cells/total T cells; MIRA, Multiplex Identification of T-cell Receptor Antigen Specificity; SARS-CoV-2, severe acute respiratory syndrome coronavirus 2; TCR, T-cell receptor.

## Discussion

Correlates of protection against COVID-19 are currently undetermined (*16*); however, cellular immunity is associated with positive outcomes (*3–5*). Here, we have provided a comprehensive assessment of the T-cell response to AZD1222 vaccination. AZD1222 vaccination elicited a robust and polyfunctional Th1-dominated T-cell response to the SARS-CoV-2 spike protein with a significant spike-specific CD8+ T-cell response across all age groups 28 days post second dose. These responses may be critical in reducing the clinical symptoms associated with COVID-19 disease, as evidenced by longitudinal follow-up of patients with SARS-CoV-2 infection, where rapid CD4+ T-cell responses in acute COVID-19 were shown to be associated with mild disease and accelerated viral clearance, and early appearance of SARS-CoV-2–specific T cells was associated with shorter duration of infection (*17*). In a study that measured SARS-CoV-2– specific antibodies, CD4+, and CD8+ T cells in participants with a range of COVID-19 disease severities, severe or fatal disease was shown to be associated with minimal or lacking SARS-CoV-2–specific CD4+ and CD8+ T-cell responses (*3*). Furthermore, SARS-CoV-2–specific CD4+ T cells were significantly associated with less severe disease. SARS-CoV-2–specific CD8+ T cells were also associated with lower disease severity, reinforcing the potential importance of T-cell–mediated immunity for minimizing disease caused by SARS-CoV-2 infection. Finally, there is evidence with some vaccinations, such as influenza, that T-cell– mediated immunity may be a more reliable correlate of protection than antibody responses in older adults (*18*). The AZD1222 primed T-cell responses that formed across all age groups may provide critical protection from severe illness due to COVID-19, as seen in vaccinated participants in these trials.

Strong Th1 responses are an important safety indicator for vaccines against respiratory pathogens. Previous whole inactivated virus vaccines for measles (*19*) and respiratory syncytial virus (*20*) led to vaccine-associated enhanced respiratory disease (VAERD). This disease, characterized by an intense allergic inflammation of the airways, is driven by vaccine-specific Th2 cells (*21*). Furthermore, vaccination of mice with SARS-CoV and Middle East respiratory syndrome coronavirus (MERS-CoV) vaccines has been shown to result in immunopathology that was attributed to Th2-biased responses (*22, 23*). Th1-predominant responses and balanced CD4+ and CD8+ T-cell responses are less likely to induce immunopathology and are therefore preferred COVID-19 vaccine characteristics. Importantly, spike-specific Th2 responses were minimal following AZD1222 vaccination in all age groups. These data are consistent with other SARS-CoV-2 vaccines, in which similar Th1-dominated responses have been reported (*6–9*).

When we further characterized this T-cell response, the majority of spike-specific Th1 and CD8+ T cells were polyfunctional across all age groups. While previous data have suggested that T cells, as analyzed by IFNγ ELISpot, do not increase following a second dose of AZD1222, the increases in polyfunctionality that are seen post second dose suggest that the quality, rather than quantity, of the response may be improved by a two-dose regimen. The quality of the T-cell response, via cytokine characterization, can give clues as to how well T cells are performing effector functions and organizing immune responses such as proliferation (*24*). Models of *Leishmania major* infection in mice have been used to understand Th1 responses to natural infection and vaccine-elicited protection (*25*). Data from these models have suggested that the magnitude of Th1 cell response was not predictive of vaccine-elicited protection. In contrast, the frequency of polyfunctional Th1 cells that simultaneously secrete IFNγ, TNF, and IL-2 correlated with pathogen control and vaccine-mediated protection (*25*). The polyfunctional responses that we observe in our analyses therefore support the contribution of T-cell immunity to the protective effect of AZD1222 vaccination. While there was more limited polyfunctionality of CD8+ T-cell responses after the first dose of AZD1222 in adults aged ≥70 years, after a second dose, CD8+ T cells improved in quantity and polyfunctionality. Therefore, two doses of AZD1222 may be required in adults aged ≥70 years to achieve the same degree of polyfunctionality that is achieved in other age groups following the first dose. Data from clinical trials and emerging real-world evidence in individuals aged ≥65 has demonstrated the effectiveness of AZD1222 for protection of COVID-19 disease following one or two doses (*13, 26–28*).

Although T cells are important for minimizing disease caused by SARS-CoV-2, nAbs have also been found to be significantly associated with protection against reinfection (*29*). B cells depend upon CD4+ T cell help for the development of pathogen-specific antibody responses (*30*). In fact, anti-spike receptor-binding domain (RBD) antibody responses in patients who have recovered from COVID-19 correlated with the magnitude of spike-specific CD4+ T-cell responses (*31*). Such antibody responses are facilitated specifically by a subset of CD4+ T cells called T follicular helper (TfH) cells via the development of germinal centers in secondary lymphoid tissues (*32*). Circulating Tfh (cTfh) cells have been detected in patients who have recovered from COVID-19 (*33*) and have even been shown to be prominent among specific CD4+ T cells in acute and convalescent COVID-19 cases (*3*). Characterizing the frequency and phenotype of cTfH cells following AZD1222 vaccination may be informative to future studies.

In the present study, AZD1222-expanded T cells were reactive to a variety of epitopes spanning the length of spike protein. Analysis by TCRseq showed an increase in both breadth and depth across multiple epitopes of the SARS-CoV-2 spike protein. Recently, the SARS-CoV-2 spike genome has accumulated mutations, resulting in the emergence of SARS-CoV-2 variants, including the B.1.351 lineage that was first identified in South Africa, and the B.1.1.7 lineage that was first identified in the UK (*34, 35*). Efficacy of AZD1222 against SARS-CoV-2 variants B.1.351 and B.1.1.7 investigated in Syrian hamsters showed that, despite an observed reduction in nAb titers against B.1.351 in AZD1222-vaccinated hamsters compared with those against B.1.1.7, there was evidence of protection in the lower respiratory tract against both variants post AZD1222 vaccination (*36*). AZD1222 vaccination has been shown to provide protection against symptomatic disease in adults aged ≥18 years caused by the B.1.1.7 lineage, shortening the duration of shedding and viral load, which may result in reduced transmission of disease (*37*). Although AZD1222 was not able to prevent mild to moderate symptomatic infection against B.1.351 in humans, we believe the vaccine may still play a protective role against severe disease and further study is warranted. Importantly, recent studies in patients exposed to SARS-CoV-2 demonstrated that CD4+ and CD8+ T-cell responses were not significantly affected by mutations found in the B.1.351 and B.1.1.7 variants (*38, 39*). Here, although nearly 30% of unique TCRs mapped to a single region of the spike protein that is mutated in the B.1.351 SARS-CoV-2 variant, it is important to note that nearly half of all unique spike-specific TCRs recognized epitopes outside of the mutated regions found in the B.1.351 and B.1.1.7 variants. Moreover, the mutation within this single region is a single point mutation, D215G. The breadth of T-cell responses across multiple viral epitopes spanning the length of SARS-CoV-2 spike following vaccination with AZD1222 suggests that the cellular immune response to the vaccine may be resilient to point changes in SARS-CoV-2.

Limitations of the study included that it was run in the UK, with a majority of participants who were white and of British descent, and with more females than males in the overall population. This may have therefore resulted in underestimating clonal diversity induced by AZD1222, due to a population with a more limited HLA profile and may be a factor responsible for reduced spike-specific CD8+ T-cell clonal diversity and lower frequencies compared with corresponding Th1 responses. Furthermore, spike-specific CD8+ T-cell frequencies may have been reduced because samples were analyzed at Day 28 post vaccination, which is 2 weeks past the peak timepoint of the T-cell responses (*13*). Lack of availability of biospecimens at this timepoint in the present study prevented analyses of AZD1222-induced T-cell responses against SARS-CoV-2 and characterization of the TCR repertoire at this peak timepoint. Furthermore, in this study, only six participants vaccinated with AZD1222 were seropositive at Day 0, providing limited data for the effect of AZD1222 vaccination on seropositive participants. Despite these study limitations, our observations are still consistent with data that SARS-CoV-2–specific CD4+ T-cell responses are more prominent than CD8+ T-cell responses following infection (*31*) and vaccination (*7, 8, 10*).

In summary, a combination of antibody and T-cell immunity is likely needed to provide long-term protection against SARS-CoV-2 infection (*40*). Previous studies have shown that AZD1222 leads to a robust nAb response across multiple age groups (*12, 13*). Here, we demonstrate that AZD1222 vaccination also induces a polyfunctional Th1-dominated T-cell response to SARS-CoV-2 spike protein in all adult age groups, including significant expansion of SARS-CoV-2 spike-specific CD4+ and CD8+ T cells, with unique TCR sequences that mapped to multiple epitope regions across the entire spike protein, which may provide long-lasting protection from severe disease associated with different SARS-CoV-2 variants.

## Materials and Methods

### Study design

Healthy adults aged 18 to ≥70 years were enrolled in a single-blind, randomized, controlled, phase 2/3 trial for the SARS-CoV-2 vaccine, AZD1222 (ChAdOx1 nCoV-19) as described in the previously published safety and immunogenicity report (*13*). Participant samples were randomly selected from three age cohorts who received either two doses of AZD1222 (5×10^10^ virus particles) or control MenACWY (fig. S1). In total, samples were obtained from 280 unique participants in studies COV001 and COV002. Full descriptions of the methods of the studies have been previously published, including full study protocols (*12, 13*). Written informed consent was obtained from all participants, and the trials were done in accordance with the principles of the Declaration of Helsinki and Good Clinical Practice. COV001 was approved in the UK by the Medicines and Healthcare products Regulatory Agency (reference 21584/0424/001-0001) and the South-Central Berkshire Research Ethics Committee (reference 20/SC/0145). COV002 was sponsored by the University of Oxford (Oxford, UK) and approved in the UK by the Medicines and Healthcare products Regulatory Agency (reference 21584/0428/001-0001) and the South-Central Berkshire Research Ethics Committee (reference 20/SC/0179). Vaccine use was authorized by Genetically Modified Organisms Safety Committees at each participating site.

Samples from 67 participants were used for ICS analysis and samples from 233 participants for TCR analysis, with 20 participant samples present in both datasets (Table 1). One patient in TCR analysis from the 18–55 age group was confirmed to have received a lower first dose (2.25×10^10^ virus particles) and a standard second dose (5×10^10^ virus particles) of AZD1222.

### ICS assay and analysis

PBMCs were isolated from blood samples within 6 hours of venipuncture and cryopreserved at ≤–150°C prior to use in ICS assays. ICS was performed on samples obtained at Day 0, Day 28, second dose (if different than Day 28), and Day 28 post second dose.

Antibody titration of all antibodies included in the analysis was performed prior to the analysis of clinical specimens. ICS assay was used to evaluate T-cell responses, as previously described (*10*). Briefly, PBMCs were thawed and rested overnight before being stimulated with pools of 15-mer peptides overlapping by 10 amino acids covering the N-terminus of SARS-CoV-2 spike protein up to the furin cleavage site (S1 pool) and the C-terminus of the SARS-CoV-2 spike protein up to the furin cleavage site (S2 pool) for 6 hours at 37°C with 5% CO_2_. Peptide pools were custom ordered from JPT and were >85% pure.

Following stimulation, cells were stained and analyzed as described previously (*41*). Briefly, cells were washed and stained with viability dye for 20 minutes at room temperature, followed by surface stain for 20 minutes at room temperature, cell fixation and permeabilization with BD cytofix/cytoperm kit (catalog # 554714) for 20 minutes at room temperature, and then intracellular stain for 20 minutes at room temperature. See Table S1 for a complete list of antibodies used. Upon completion of staining, cells were collected on a BD FACSymphony Flow Cytometer. Samples were invalidated if <20,000 live CD3+ T cells were collected. On average, ≥175,000 were collected for each sample. Samples were analyzed using FlowJo 10.6.2. Anomalous “bad” events were separated from “good” events using FlowAI (*42*). “Good events” were used for all downstream gating. Cytokine positive cells were determined by gating indicated in fig. S5. Individual cytokines were plotted on the Y-axis versus CD69+ cells on the X-axis, and only CD69+ events were used to determine positive responses. No cytokine-positive responses were detected above background in the CD69-gate. CD69+ activated cells were increased over baseline following AZD1222 vaccination but did not significantly differ between peptide pools or by age groups (fig. S6). All antigen-specific cytokine frequencies are reported after background subtraction of identical gates from the same sample incubated with negative control stimulation (DMSO).

Assay qualification was performed by the Vaccine Immunology Program at the Vaccine Research Center, National Institute of Allergy and Infectious Diseases, Gaithersburg, MD, USA. Assay qualification included assessment of Th1 and Th2 specificity, Th1 and Th2 intra-assay precision, inter-assay precision, and SARS-CoV-2 peptide pool validation. Th1 and Th2 specificity was conducted by utilizing PBMCs from acutely infected cytomegalovirus (CMV) donors (for Th1 specificity) or *Filaria* parasite donors (for Th2 specificity), given the established cytokine profiles of these pathogens.

### T-cell receptor β chain sequencing

Immunosequencing of the CDR3 regions of human TCRβ chains was performed using the ImmunoSEQ^®^ Assay (Adaptive Biotechnologies, Seattle, WA). Extracted genomic DNA was amplified in a bias-controlled multiplex polymerase chain reaction (PCR), followed by high-throughput sequencing. Sequences were collapsed and filtered to identify and quantitate the absolute abundance of each unique TCRβ CDR3 region for further analysis as previously described (*43–45*). The fraction of T cells was calculated by normalizing TCRβ template counts to the total amount of DNA usable for TCR sequencing, where the amount of usable DNA was determined by PCR amplification and sequencing of several reference genes that are expected to be present in all nucleated cells.

### Mapping of SARS-CoV-2 TCRβ sequences

TCR sequences from participants receiving AZD1222 were mapped against a set of TCR sequences that are known to react to SARS-CoV-2. Briefly, these sequences were first identified by MIRA (*46*). TCRs that reacted were further screened for enrichment in COVID-19–positive repertoires collected as part of immuneCODE (*15*) and compared with COVID-19–negative repertoires to remove TCRs that may be highly public or cross-reactive to common antigens. Individual response could be quantified by the number and/or frequency of SARS-CoV-2 TCRs seen post vaccine. TCRs were further analyzed at the level specific open reading frame (ORF) or position within spike based on the MIRA antigens.

### Comparison with known variants

Sequences of known variants were obtained from GISAID (www.gisaid.org) and aligned to known MIRA antigen locations. Antigens that contain any mutations observed in the B.1.1.7 or B.1.351 variants were labeled as potentially impacted.

## Data Availability

TCRβ immunosequencing data that support the findings of this study have been deposited at the publicly available immuneACCESS platform at: https://clients.adaptivebiotech.com/pub/

## Acknowledgments

Writing and editing assistance, including revision of drafts under the direction and guidance of the authors, incorporating author feedback, and manuscript submission, was provided by Heather Shawcross, Emily Feist, and Rebecca Lawson (Fishawack Health). This support was funded by AstraZeneca.

## Author contributions

### Competing interests

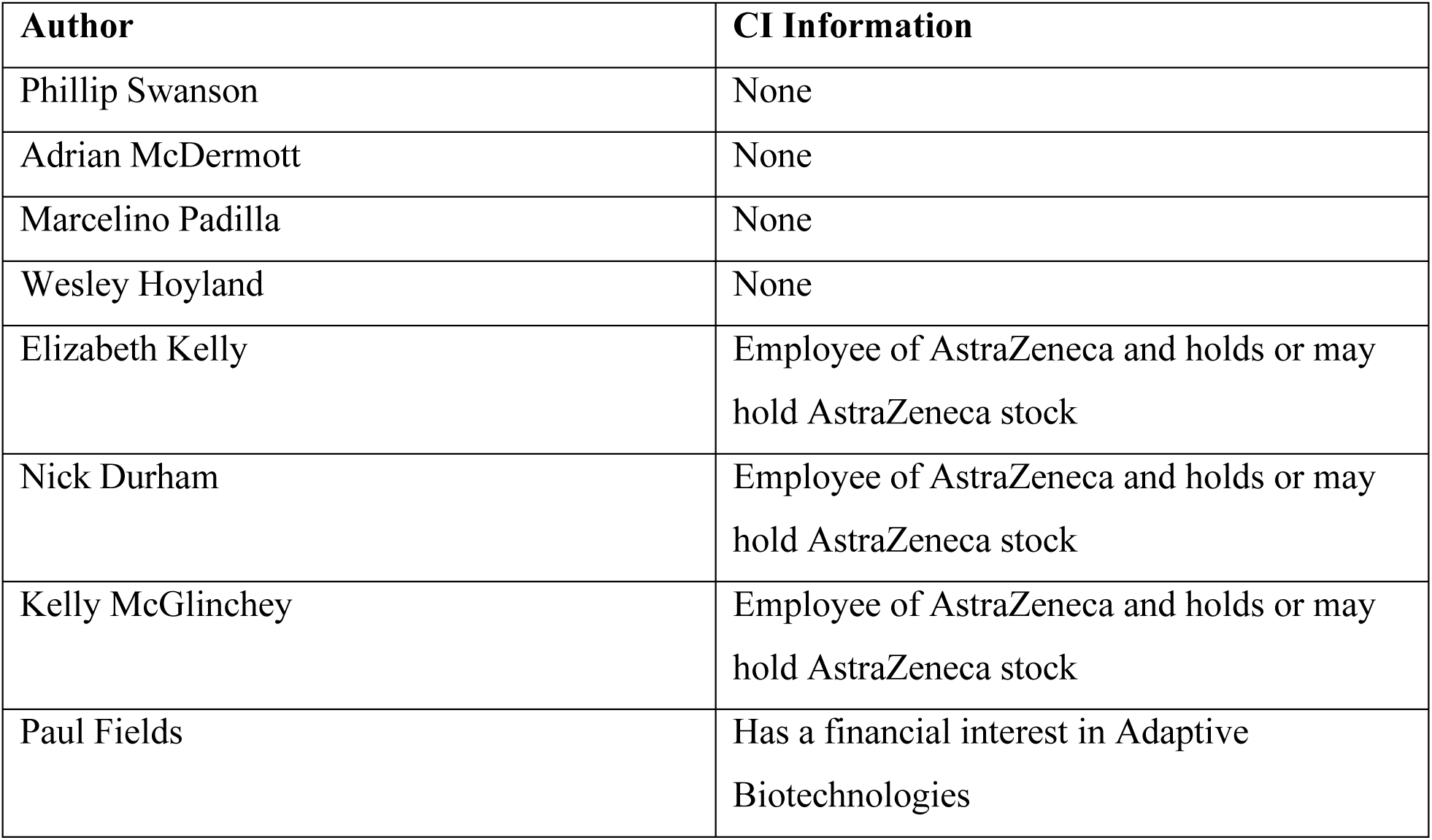
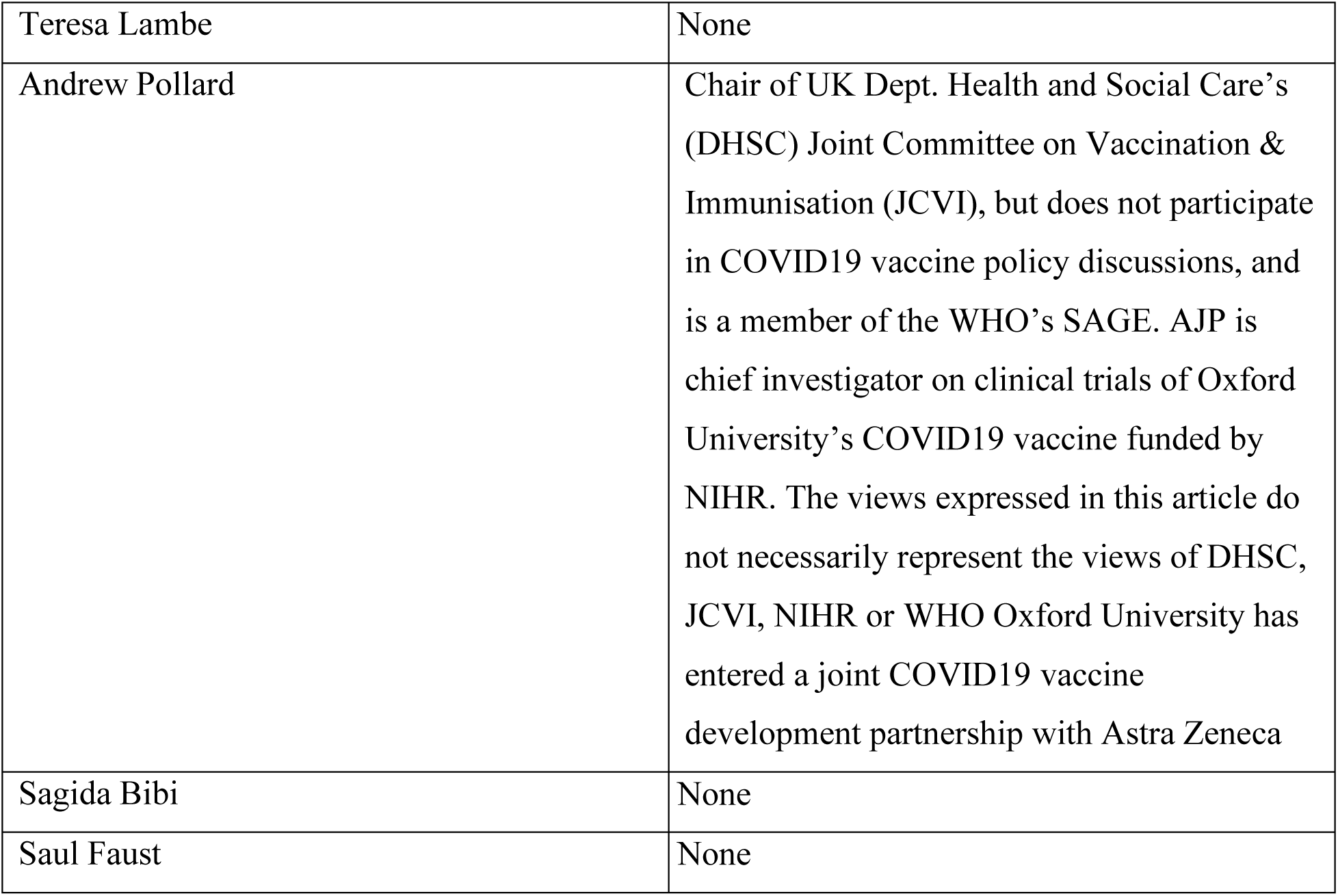

## Data and materials availability

TCRβ immunosequencing data that support the findings of this study have been deposited at the publicly available immuneACCESS® platform at: https://clients.adaptivebiotech.com/pub/

