## Supplemental Table and Figures for "T-cell mediated immunity after AZD1222 vaccination: A polyfunctional spike-specific Th1 response with a diverse TCR repertoire"

### Supplementary Materials

#### Table S1. List of antibodies used in intracellular cytokine staining

| Detector | Fluorophore | Specificity | Clone | Dilution | Stain |
|---|---|---|---|---|---|
| B710 | BB700 | IL-4 | MP4-25D2 | 1:160 | Intracellular |
| G560 | PE | CD28 | CD28.2 | 1:20 | Surface |
| G610 | ECD | CD69 | TP1.55.3 | 1:20 | Intracellular |
| G660 | PE/Cy5 | CD8a | RPA-T8 | 1:40 | Surface |
| G780 | PE/Cy7 | IFNγ | B27 | 1:160 | Intracellular |
| R660 | APC | IL-2 | MQ1-17H12 | 1:20 | Intracellular |
| R780 | APC/H7 | CD3 | SK7 | 1:80 | Intracellular |
| U395 | BUV395 | CCR7 | 150503 | 1:40 | Surface |
| U450 | UV-Blue | Viability | - | 1:500 | Surface |
| U570 | BUV563 | TNF | Mab11 | 1:10 | Intracellular |
| U785 | BUV805 | CD4 | SK3 | 1:10 | Surface |
| V450 | BV421 | IL-13 | JES 10-SA2 | 1:10 | Intracellular |
| V785 | BV785 | CD45RO | UCHL1 | 1:20 | Surface |

**Fig. S1. T-cell responses to individual peptide pools following vaccination with AZD1222 or MenACWY**. To determine spike-specific responses, PBMCs were stimulated with two different peptide pools covering the entire SARS-CoV-2 spike protein (S1 and S2 pools). The responses for both pools were combined for Fig. 1 to determine the overall spike-specific response. Shown here are the data from Fig. 1, deconstructed into the T-cell responses to each individual peptide pool. Th1 (A), Th2 (B), and CD8 (C) T-cell cytokine responses following stimulation with the indicated peptide pool from AZD1222 (left) or MenACWY (right) vaccinated participants. In the box and whisker plots, the horizontal line represents median, boxes represent interquartile range, whiskers extend to 5th and 95th percentiles, and symbols represent outlier samples. MenACWY, meningococcal conjugate vaccine; PBMCs, peripheral blood mononuclear cells; SARS-CoV-2, severe acute respiratory syndrome coronavirus 2; Th1, T cell helper type 1.

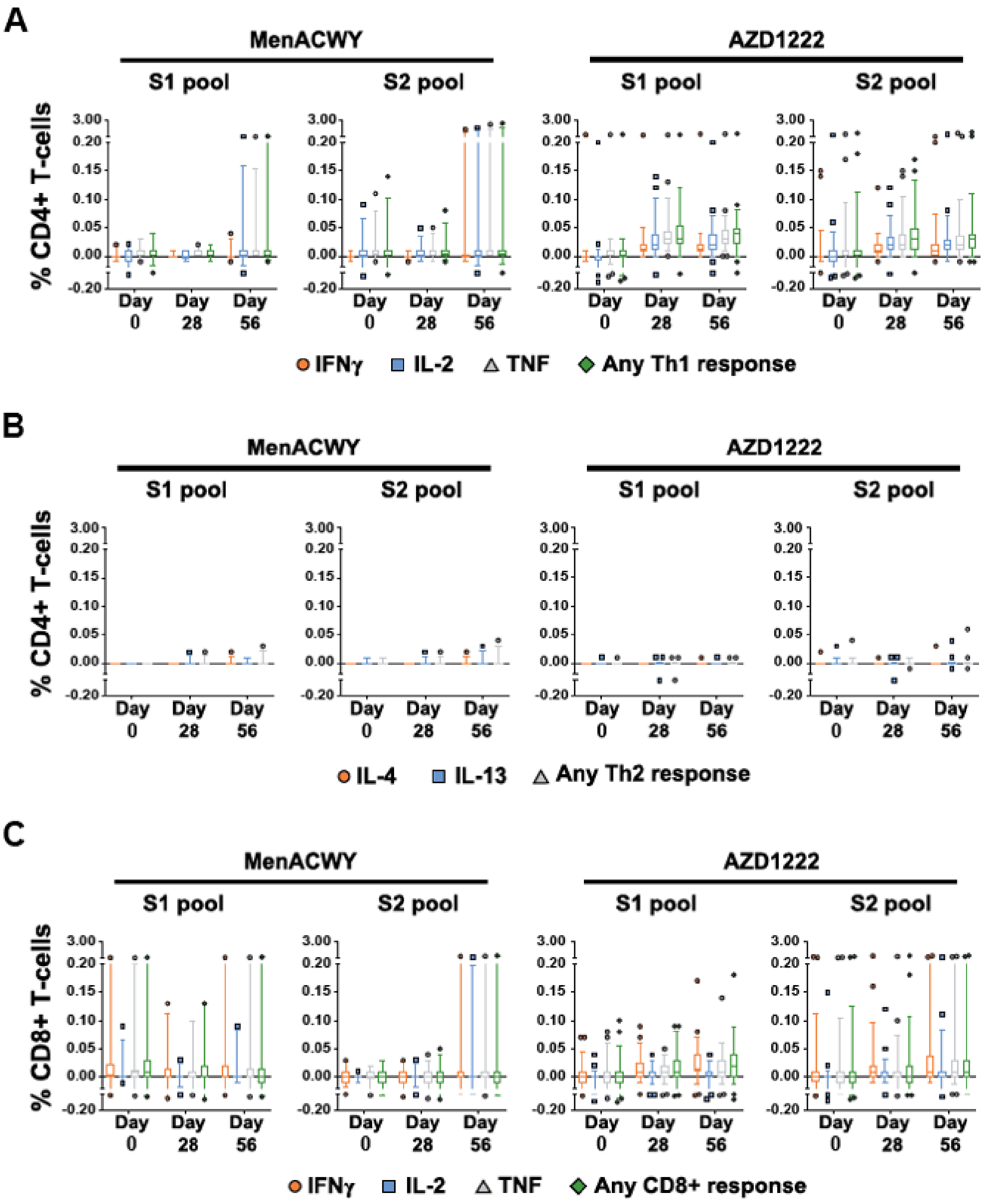

**Fig. S2. Individual age-specific T-cell responses to AZD1222 vaccination.** Frequencies of CD4 T cells (A) or CD8 T cells (B) from participants within each age cohort producing IFNγ, IL-2, TNF, or any combination of these cytokines at the indicated timepoints following stimulation with SARS-CoV-2 spike peptide pools. SARS-CoV-2, severe acute respiratory syndrome coronavirus 2; Th1, T cell helper type 1.

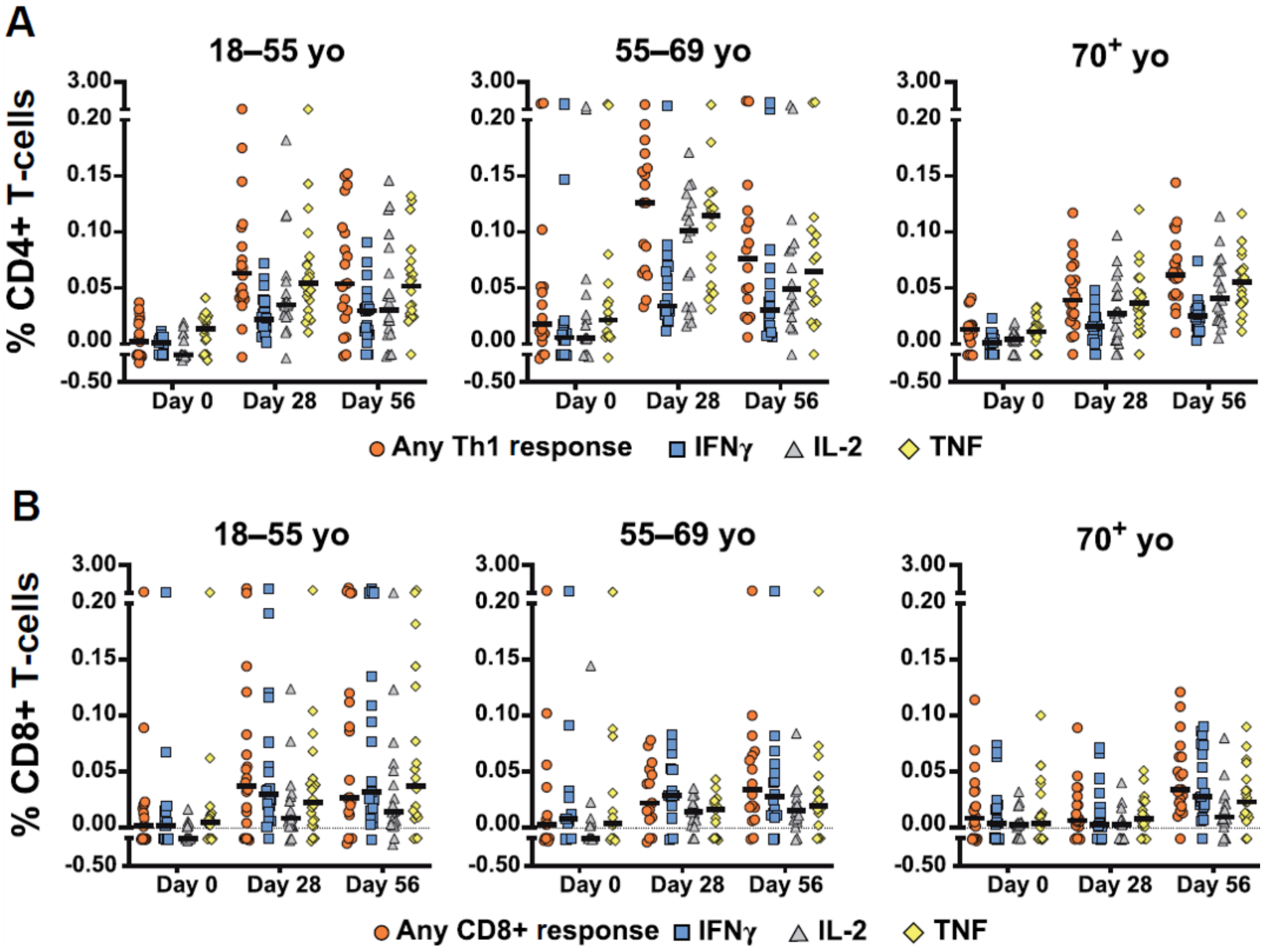

**Fig. S3. Spike-specific TCR depth and breadth following vaccination with AZD1222 at 4-week and 12-week second dose schedules.** Spike-specific TCR depth (A) and breadth (B) following vaccination with AZD1222 at approximately 4-week (18–60 days) and approximately 12-week (61–130 days) second dose schedules. In the box and whisker plots, the horizontal line represents median, boxes represent interquartile range, whiskers extend to 5th and 95th percentiles, and symbols represent outlier samples. Significant differences determined by Sidak's multiple comparisons tests. ns, not significant, ****p<0.0001. Breadth, SARS-CoV-2 associated unique TCRs/total unique TCRs; D2, Dose 2; Depth, SARS-CoV-2 associated T cells/total T cells; MenACWY, meningococcal conjugate vaccine; PB, post booster; SARS-CoV-2, severe acute respiratory syndrome coronavirus 2; TCR, T-cell receptor.

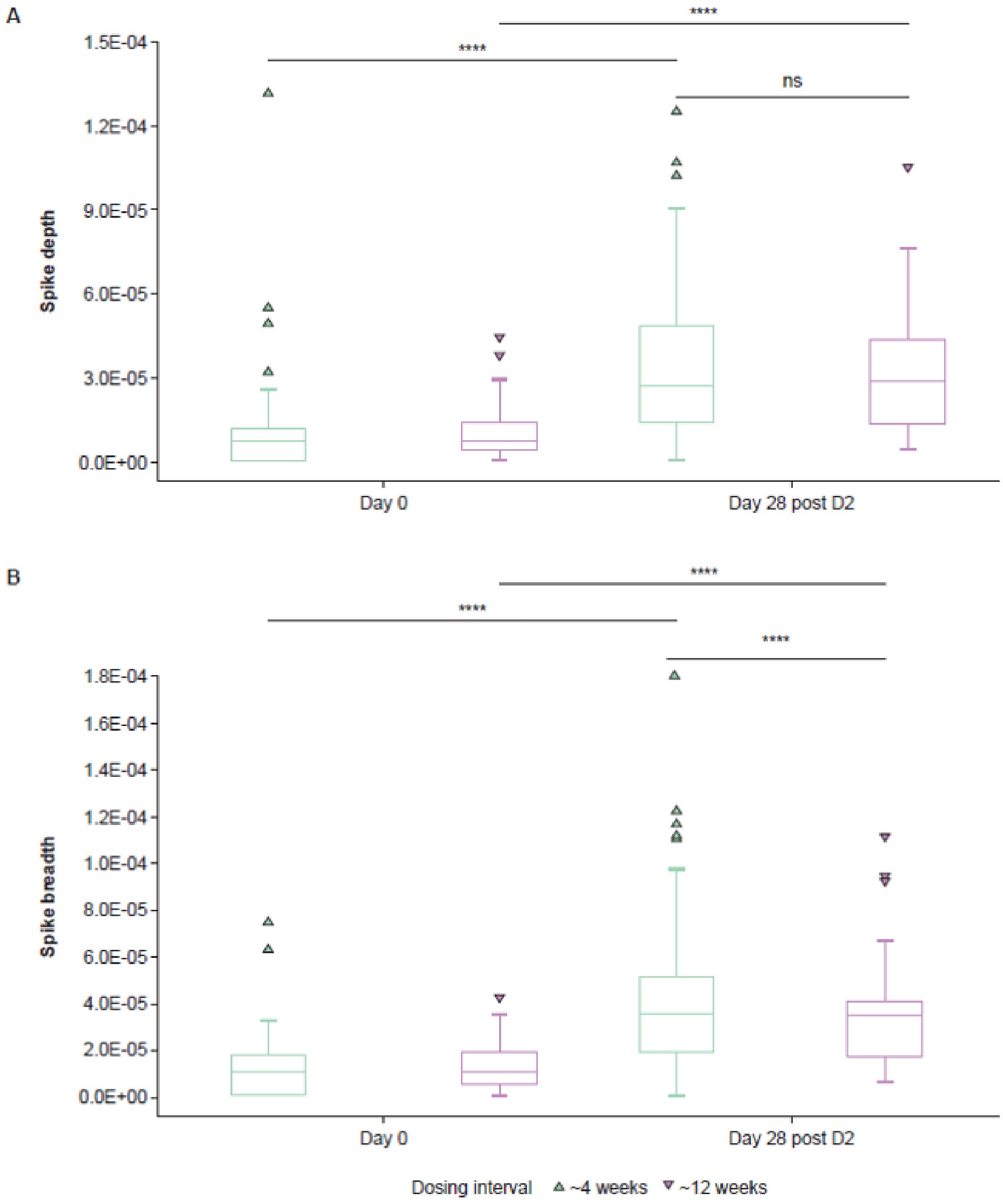

**Fig. S4. Non-spike-specific TCR breadth and depth following vaccination with AZD1222 or MenACWY.** (A) Non-spike-specific TCR breadth following vaccination with MenACWY (blue) or AZD1222 (orange). (B) Non–spike-specific TCR depth following vaccination with MenACWY (blue) or AZD1222 (orange). In the box and whisker plots, the horizontal line represents median, boxes represent interquartile range, whiskers extend to 5th and 95th percentiles, and symbols represent outlier samples. Significant differences determined by Sidak's multiple comparisons tests. ns, not significant. Breadth, SARS-CoV-2 associated unique TCRs/total unique TCRs; D2, Dose 2; Depth, SARS-CoV-2 associated T cells/total T cells; MenACWY, meningococcal conjugate vaccine; SARS-CoV-2, severe acute respiratory syndrome coronavirus 2; TCR, T-cell receptor.

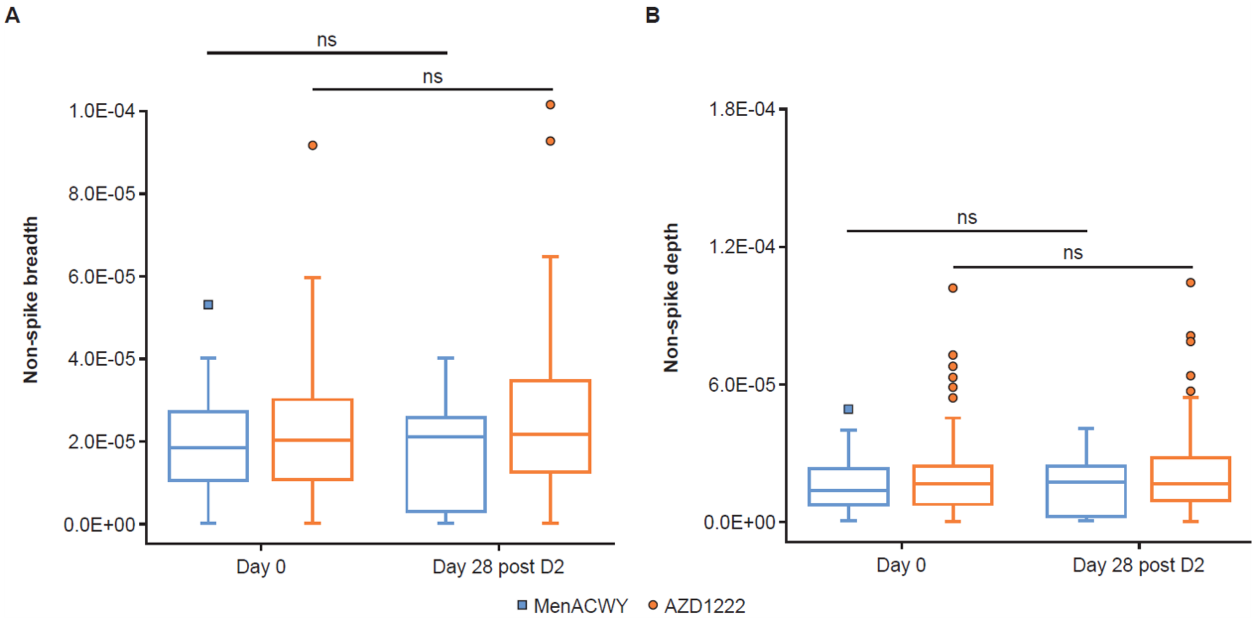

**Fig. S5. Flow cytometric gating strategy.** PBMCs stained with the 13-color panel. (A) Lineage gating of T cells. After gating on single cells, lymphocytes, and viable CD3+ T cells, T cells were further subdivided into CD4+ and CD8+ T cells. (B,C) Gating to measure T-cell function following no stimulation or stimulation with SARS-CoV-2 S1 peptide pool. Cytokine staining is shown following gating on CD4 T cells (B) or CD8 T cells (C). Memory T-cell subset gating for CD4 T cells (D) and CD8 T cells (E). T-cell populations identified include Naïve (Tn), central memory (Tcm), transitional memory (Ttm), effector memory (Tem), and terminal effector (Tte) cells. Proportion of polycytokine responses are represented by the black (all three cytokines) and gray (two cytokines) arcs. PBMCs, peripheral blood mononuclear cells, SARS-CoV-2, severe acute respiratory syndrome coronavirus 2.

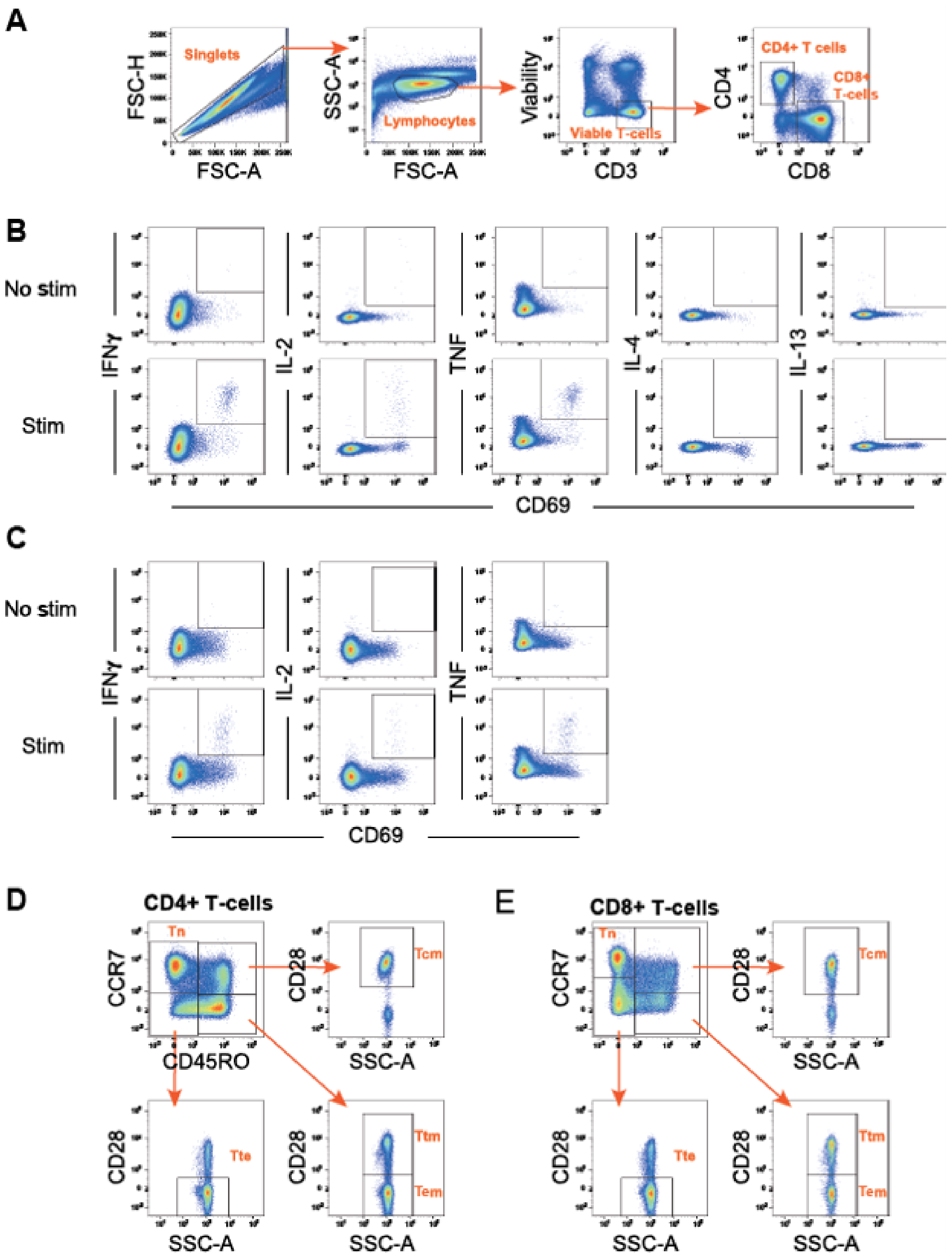

**Fig. S6. T-cell activation state.** Frequencies of spike-specific (cytokine$^{+}$) and non-specific (cytokine$^{-}$) CD4 (A) and CD8 (B) T cells expressing CD69 were measured. T-cell frequencies following stimulation with S1 (left) and S2 (right) peptide pools for participants in each age group at the indicated timepoints are shown. In the box and whisker plots, the horizontal line represents median, boxes represent interquartile range, whiskers extend to 5th and 95th percentiles, and symbols represent outlier samples.

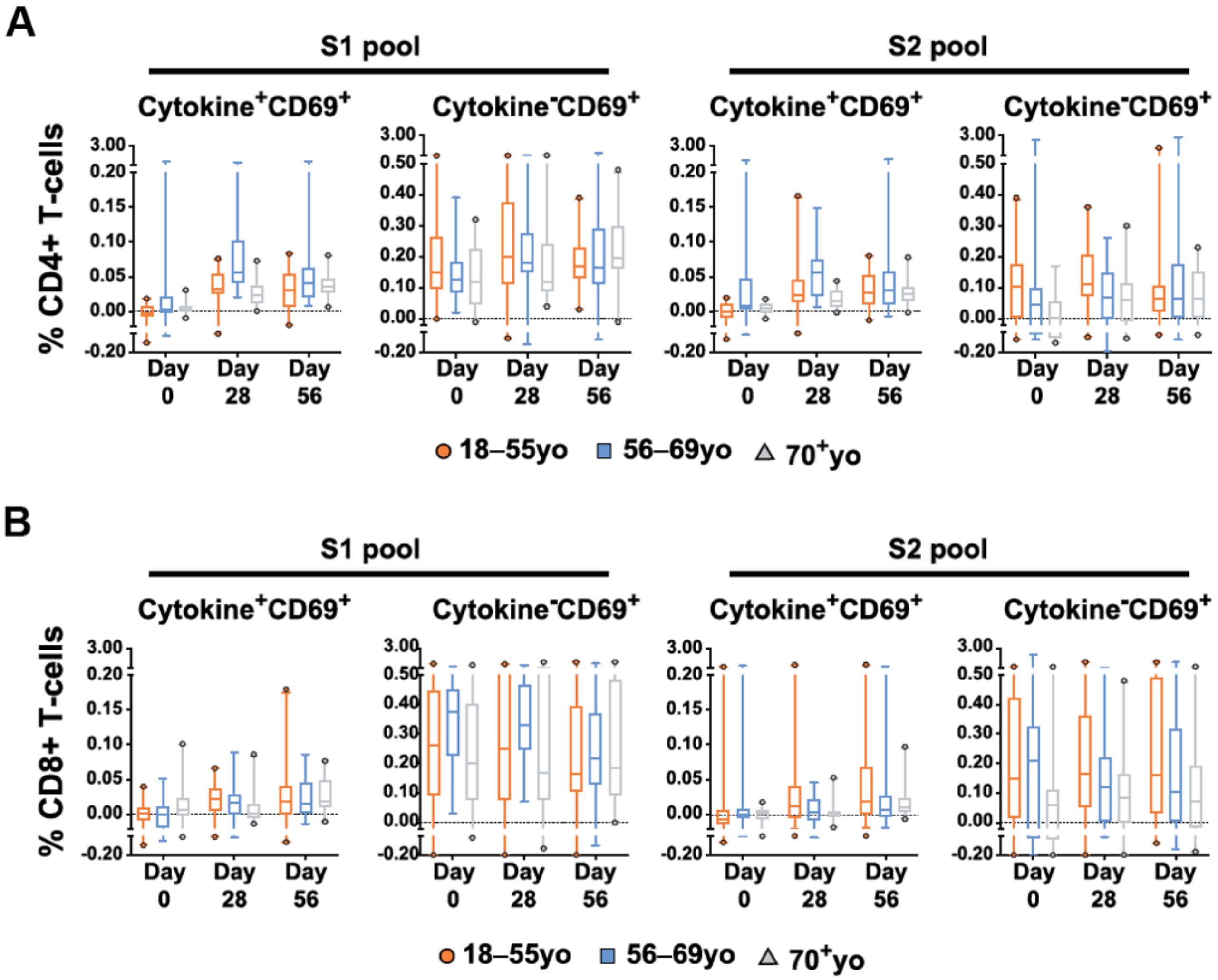
